# Mitochondrial DNA-Mediated Immune Activation After Resuscitation from Cardiac Arrest

**DOI:** 10.1101/2025.02.14.25322318

**Authors:** Tyler J. Rolland, Emily R. Hudson, Luke A Graser, Sumbule Zahra, Daniel Cucinotta, Swati D. Sonkawade, Umesh C. Sharma, Brian R. Weil

## Abstract

**Background:** Post-cardiac arrest syndrome (PCAS) is characterized by a robust inflammatory response that contributes to significant morbidity and mortality among patients resuscitated from sudden cardiac arrest (SCA). Mitochondrial DNA (mtDNA), with its bacterial-like genomic motifs, has been implicated as a damage-associated molecular pattern in other inflammatory contexts, but its role as a pro-inflammatory stimulus in PCAS has not been studied. Accordingly, the present study was designed to determine if PCAS is characterized by a rise in circulating mtDNA and, if so, whether mtDNA is selectively released, how it activates immune cells, and if targeting mtDNA-sensing pathways attenuates leukocyte activation.

**Methods:** Plasma mtDNA and nuclear DNA (nucDNA) levels were measured in peripheral blood samples collected ∼4-hours post-ROSC from swine with PCAS (n=8) and patients hospitalized after resuscitation from out-of-hospital cardiac arrest (OHCA; n= 57). Additionally, *in vitro* studies were performed where porcine peripheral blood mononuclear cells (PBMCs) were treated with mtDNA or extracellular vesicles (EVs) isolated from post-ROSC plasma. Pharmacological inhibitors were utilized to inhibit toll-like receptor 9 (TLR9)- and cyclic GMP–AMP synthase (cGAS)-mediated mtDNA sensing.

**Results:** A significant ∼250-fold elevation in circulating mtDNA was observed shortly after ROSC in swine despite negligible changes in circulating nucDNA, suggesting selective release of mtDNA in PCAS. This finding was corroborated in human OHCA survivors, in which circulating mtDNA was similarly elevated during the early post-ROSC period. Circulating mtDNA was largely encapsulated within EVs in swine and humans, suggesting a conserved mechanism of release across species. *In vitro* studies demonstrated that PBMC internalization of mtDNA-containing-EVs was required for immune activation and promoted development of a pro-inflammatory leukocyte phenotype characterized by altered surface marker expression and increased release of TNFα, IL-1β, and IL-6. Disrupting EVs or degrading enclosed DNA attenuated these responses, which were partially restored upon reintroduction of mtDNA. Pharmacological blockade of TLR9 or cGAS pathways significantly reduced mtDNA-induced inflammation, providing insight regarding signaling pathways that may be targeted to modulate mtDNA-mediated immune activation in PCAS.

**Conclusions:** These novel findings demonstrate that brief whole-body ischemia and reperfusion in the context of resuscitation from SCA triggers selective mtDNA release, primarily within EVs, that acts as a potent driver of immune activation in PCAS. By linking EV-encapsulated mtDNA to TLR9 and cGAS activation, this study provides a foundation for the development of novel therapeutic interventions aimed at limiting mtDNA release or disrupting its downstream sensing pathways to enhance survival and improve outcomes after SCA.

**Clinical Perspective:** *What is new?:*

- Our study reveals that circulating mitochondrial DNA (mtDNA), primarily encapsulated in extracellular vesicles (EV), is selectively released into the bloodstream after resuscitation from sudden cardiac arrest.
- EV-encapsulated mtDNA triggers immune cell activation, evidenced by phenotypic shifts toward inflammatory dendritic cells and macrophages, as well as increased pro-inflammatory cytokine secretion.
- Pharmacological inhibition of TLR9 and cGAS pathways significantly attenuates the mtDNA-induced inflammatory response, pointing to novel therapeutic avenues for modulating post-resuscitation immune activation in patients with post-cardiac arrest syndrome (PCAS).

*What are the clinical implications?:*

- Identification of mtDNA as a key driver of sterile inflammation in PCAS highlights a potential target for interventions aimed at reducing multi-organ damage and improving neurological outcomes.
- Therapeutic strategies to block mtDNA release or downstream signaling (e.g., TLR9/cGAS inhibition) may limit harmful pro-inflammatory cascades and bolster long-term survival following resuscitation from cardiac arrest.
- Early clinical screening for elevated EV-encapsulated mtDNA could help refine prognostic evaluations, complement current biomarkers, and guide personalized therapy to lessen the inflammatory burden of PCAS.

## INTRODUCTION

Each year in the United States, nearly 600,000 adults experience sudden cardiac arrest (SCA), often with little warning and frequently outside the hospital^1^. Although improvements in cardiopulmonary resuscitation have increased initial survival rates, fewer than 10% of these patients regain normal brain function and survive long-term^2,3^. A primary reason for these dismal post-resuscitation outcomes is post-cardiac arrest syndrome (PCAS), a condition characterized by systemic inflammation, cardiac dysfunction, and neurological injury following return of spontaneous circulation (ROSC)^4,5^. Higher levels of inflammatory cytokines (e.g., TNFα, IL-1β, IL-6) are linked to poor outcomes in cardiac arrest survivors, emphasizing the possible clinical impact of reducing unnecessary immune activation^6,7^. Unfortunately, the fundamental mechanisms underlying the robust post-resuscitation inflammatory response remain poorly understood, which has hindered the development of effective treatments for PCAS.

Recent findings indicate that innate immune activation triggered by danger-associated molecular patterns (DAMPs) is central to PCAS severity, but specific pro-inflammatory stimuli have yet to be identified^8^. One intriguing candidate is mitochondrial DNA (mtDNA). Because mitochondria descended from α-proteobacteria^9^, mtDNA retains bacteria-like features, such as unmethylated CpG repeats, that can provoke an immune response^10,11^. Clinical and preclinical data suggest a link between elevated mtDNA levels and inflammation in a variety of contexts including heart failure^12^, atherogenesis^13^, and ischemic heart disease. For example, in experimental myocardial infarction^14^ and clinical cardiopulmonary bypass^15^, elevated circulating mtDNA levels correlate with increased circulating and tissue levels of inflammatory cytokines. Preliminary work also suggests that mtDNA can accumulate in the bloodstream after oxidative stress without overt necrosis^6,16^, potentially acting as a powerful trigger for sterile inflammation. However, whether mtDNA is selectively released after brief whole-body ischemia in the context of SCA has not been directly studied. Moreover, how circulating mtDNA might be detected by leukocytes (via Toll-like receptor 9 (TLR9)^17^ or the cyclic GMP-AMP synthase–stimulator of interferon genes (cGAS-STING) pathway^18^) and whether blocking these pathways can dampen post-resuscitation immune activation is unknown.

Accordingly, we aimed to delineate the role of mtDNA in activating immune cells after SCA and determine whether interfering with mtDNA-mediated inflammatory signaling pathways could mitigate the pro-inflammatory leukocyte activation. By integrating clinical data from out-of-hospital cardiac arrest survivors with mechanistic experiments in porcine models of systemic (SCA) and regional brief ischemia (BI), we tested the hypothesis that selective release of mtDNA, rather than nuclear DNA (nucDNA), triggers robust post-ROSC immune activation via TLR9 and cGAS signaling pathways. As detailed in the results below, our results show that extracellular vesicle (EV)-encapsulated mtDNA serves as a potent inflammatory stimulus in PCAS, offering a novel therapeutic avenue to potentially improve outcomes among patients resuscitated from SCA.

## METHODS

### Data Availability

The data supporting the study’s findings are available from the corresponding author upon reasonable request. Please see **Supplemental Materials** for complete methodological details.

### Ethics Statement

Investigators were blinded to each patient and animal’s experimental group (e.g., baseline vs. 1-hr post-ROSC vs. 4-hr post-ROSC) while performing blood sampling, *in vitro* experimentation, and data analysis. All experimental procedures and protocols conformed to institutional guidelines for the care and use of animals in research and were approved by the State University of New York at Buffalo Institutional Animal Care and Use Committee (ID: MED15083Y) and Institutional Review Board (ID: STUDY00000284).

### Porcine SCA Model

A total of eight Yorkshire-cross farm-bred pigs (2M/6F) were studied in the closed-chest state using a protocol summarized in **Figure S1**, as described previously^19^. Briefly, animals were subjected to 10-minutes of cardiac arrest, followed by up to 20-minutes of cardiopulmonary resuscitation (CPR) with manual chest compressions, mechanical ventilation, and defibrillation until achieving return of spontaneous circulation (ROSC; unassisted arterial systolic blood pressure (SBP) of ≥ 80 mmHg for at least 1 minute). If ROSC was achieved, blood plasma samples were collected at 1- and 4-hr post-ROSC.

### Porcine Brief Myocardial Ischemia (BI) Model

A total of five male Yorkshire-cross farm-bred pigs were studied in the closed-chest state to induce a transient 10-minute occlusion of the left anterior descending (LAD) coronary artery, as described previously^20^. In short, animals were subjected to 10-minutes of brief ischemia (BI) localized to the LAD perfusion territory via inflation of a percutaneous angioplasty balloon. After 10-minutes of ischemia, the balloon was deflated, blood flow was restored, and subjects were survived for 1-hour post-reperfusion (post-BI). Plasma samples from the coronary sinus (CS) were collected before the localized ischemic event (pre-BI) and 1-hour post-BI (full methodological details provided in the **Supplemental Materials**).

### Human Out-of-Hospital Cardiac Arrest (OHCA) Plasma Sample Collection

In a prospective cohort study, 57 successfully resuscitated patients of non-traumatic OHCA were recruited from four tertiary-level hospitals in Buffalo, Western New York, and followed for 30 days following ROSC. Patients with active cancer, infection, cardiac pathology, or surgery in the three months prior to presentation were excluded. Cardiac arrest characteristics, demographics, comorbidities, and therapeutic interventions were recorded (**Table S4)**, and plasma samples were collected between 2-6 hours following ROSC and frozen at −80°C to be analyzed via qPCR. OHCA samples were compared against healthy control subjects.

### Plasma mtDNA and nucDNA Digestion and EV Lysis

To digest circular mtDNA and nucDNA from plasma and plasma-EVs, samples were treated with 50U/mL DNase I (Thermo, Ref: 89836) at 37°C for 1 hour. Subsequently, DNase I was inactivated with 50mM EDTA for 10 minutes at 65°C, per the manufacturer’s protocol. To lyse open EVs, 1x RIPA buffer was added to samples and incubated on ice for 30 minutes.

### Quantification of Plasma mtDNA, nucDNA, and Total dsDNA via qPCR

Whole plasma from patients and swine was clarified by centrifugation of 500xg for 5-minutes before transferring the supernatant to a DNeasy Blood and Tissue Kit (Qiagen, Ref: 69504) to isolate plasma DNA, per the manufacturer’s protocol. To identify EV-enclosed DNA, isolated plasma-EVs were additionally treated with DNase I (described above) to remove free-floating and non-encapsulated DNA before isolation via a DNeasy kit. Following DNA isolation, human and porcine samples were quantified by qPCR using SsoAdvanced Universal SYBR Green Supermix (BIO-RAD, Ref: 1725270) against a standard curve of mtDNA and nucDNA in a known concentration using primers to detect mitochondrial (Cyt-B, D-Loop) or nuclear (β2M) genes (**Table S1**), as previously described^21^. Additionally, non-specific quantification of double-stranded DNA (dsDNA) was conducted using Quanti-iT PicoGreen dsDNA Assay Kit (Thermo, Ref P7589), per the manufacturer’s protocol.

### qPCR of Inflammatory Transcripts

RNA was extracted from thawed cell lysate via RNeasy Kit (Qiagen, Ref: 74104), per the manufacturer’s protocol. A NanoDrop spectrophotometer (Thermo Scientific, Ref: ND2000) was used to determine RNA purity and concentration. Total RNA was reverse transcribed and analyzed by qPCR as previously described^22^. SsoAdvanced Universal SYBR Green Supermix (BIO-RAD, Ref: 1725270) was used in combination with primers for IFNα, IL-1α, IL-1β, IL-6, IL-8, TNFα, and beta-2-microglobulin (β2M) (**Table S1**). The comparative cycle threshold method was used to determine the mRNA expression for each target gene using the gene for β2M as the reference.

### Quantification of Inflammatory Cytokine Release, ROS/RNS, and Intracellular cGAMP

Conditioned media (CM) was collected 24-hours post-activation from the supernatant of each cell culture dish well. CM samples were clarified by centrifugation at 500xg for 5-minutes to remove cellular debris. CM was thawed and quantified in duplicate with porcine-specific ELISA kits according to manufacturer’s instructions (**Table S2**). Additionally, CM levels of reactive oxygen and nitrogen species (ROS/RNS) were measured using a DCF-based kit (Abcam, Ref: ab238535), while intracellular levels of 2’3’-cyclic guanosine monophosphate–adenosine monophosphate (cGAMP; Invitrogen, Ref: EIAGAMP) after a 30-minute incubation with 1x RIPA buffer on ice, following the manufacturer’s recommendations.

### PBMC Isolation and Culture

Naïve peripheral blood mononuclear cells (PBMCs) were isolated from healthy White Yorkshire x Landrace swine (Oak Hill Genetics, Ewing, IL). Approximately 8mL of whole blood was collected and added to Ficoll CPT Vacutainers (BD, Ref: 362761), followed by centrifugation at 1,500xg for 28-minutes at 4°C. As per the manufacturer’s protocol, the PBMC layer was collected, filtered through a 40µm filter into a 50mL Falcon tube, and washed with 20mL of PBS. The filtered PBMCs were then centrifuged at 500xg for 5-minutes and washed with PBS. Before plating, isolated PBMCs were treated with red blood cell lysis buffer for 5-minutes at room temperature, followed by centrifugation and PBS washing. Purified PBMCs were resuspended in RPMI 1640 supplemented with 1x penicillin/streptomycin, 1x sodium pyruvate, and 10% fetal bovine serum (FBS). The cells were cultured at 37°C in a humidified atmosphere of 5% CO2 and ambient oxygen for one week, with bi-daily media changes. After one week, the cells were subjected to activation assays. After aspiration of the culture medium and washing with PBS, naïve PBMCs were activated for 24 hours with either 1× LPS (5µM; Invitrogen, Ref: 00-4976-93) or 2μg/mL mtDNA with Lipofectamine 2000 Transfection Reagent (TR; Invitrogen, Ref: 11668027), as previously described^23^. During this activation period, pre- and post-ROSC plasma-EV samples were labeled with 5µM Vybrant DiD Lipophillic Dye (Thermo, Ref: V22887) and were added in place of FBS. At the end of the 24-hour stimulation period, the conditioned medium was harvested and stored at −20°C for subsequent ELISA and ROS/RNS assays, while the cells were collected for flow cytometric analysis or lysed in Qiazol Reagent for downstream qPCR. Each figure represents the collection of biologically distinct PBMC populations, therefore leading to minute differences between experimental results.

### Small Molecule Inhibition of mtDNA-Induced PBMC Inflammation

To mimic the conditions under which a therapeutic agent would be delivered to successfully resuscitated patients, small molecular inhibitions were added in suspension to basal media at the same time as activation by mtDNA + TR. Inhibitors were utilized in a concentration-response assay against TLR9 (2-8µM ODN 2088; InvivoGen, Ref: tlrl-2088 or 2-8µM ODN inh-18; InvivoGen, Ref: tlrl-inh18), cGAS (5-20µM G140; InvivoGen, Ref inh-g140), STING (1-4µM H151; InvivoGen, Ref: inh-h151), and PKG1 (1-4µM Rp-8-Br-PET-cGMP; Tocris, Ref: 3028), as previously described^24–26^. All small molecular inhibitors were prepared and frozen at −20°C until being thawed immediately before use.

### Plasma EV Isolation

Plasma from humans or swine was centrifuged at 500xg for 5-minutes to remove large particulates. The resulting supernatant was filtered through a 0.2µm filter to eliminate residual debris. Further purification was achieved using an Amicon® Ultra Centrifugal Filter (100kDa MWCO), with centrifugation at 4,000g for 60-minutes at 4°C. The isolated plasma-EV were then diluted in ratios of 1:1,000 and characterized using a ZetaView® x20 Series (Particle Metrix) particle tracking analysis. Additionally, the plasma-EV preparations were evaluated using the Exo-Check™ Exosome Antibody Array (SBI, Ref: EXORAY200), according to the manufacturer’s instructions.

### Leukocyte Isolation and Flow Cytometry

Following a 24-hour activation period, cultured leukocytes were labeled for porcine-specific inflammatory cell-surface markers ^27,28^ and viability was assessed by 7AAD staining. Flow cytometric assessment was performed on a BD LSRFortessa™ Cell Analyzer and analyzed via De Novo Software FCS Express 7 Plus (version 7.16.0035) following consistent gating strategy implementation (**Figure S2)** to quantify sub-populations of inflammatory granulocytes (CD172+CD163-), inflammatory dendritic cells (CD172+CD16+), and inflammatory macrophages (CD14+CD163+) ^27,28^ (**Table S3**). Additional staining of plasma-EVs with 5µM Vybrant DiD Lipophillic Dye allowed the detection of plasma-EV uptake by PBMC populations

### Statistical Analysis

Data are reported as mean ± standard error of the mean (SEM) or median [IQR1, IQR3], as determined by Shapiro-Wilk Tests to assess normality. Between-group differences in endpoints measured across multiple samples were assessed by repeated measures ANOVA and the post-hoc Tukey HSD test, while between-group differences in endpoints of two samples were assessed by two-tailed unpaired Student’s t test or two-tailed Mann Whitney U-Test where appropriate. For concentration-dependent inhibition curves, simple linear regressions were utilized to determine the 50% inhibitory concentration (IC50) and the coefficient of determination (R2). Significance was set to an alpha level of 0.05. All data were compiled with Microsoft Excel (version 2402), while analysis and plotting were performed with GraphPad Prism Software (version 10.2.0).

## RESULTS

### Selective Elevation of Circulating mtDNA in Pigs and Humans with PCAS

To investigate the relevance of mtDNA release in clinical PCAS, we measured circulating mtDNA levels in successfully resuscitated out-of-hospital cardiac arrest (sR-OHCA) patients (n=57; **Table S4**) within 6-hours of ROSC. Relative to healthy controls (n=9), sR-OHCA patients exhibited significantly elevated plasma mtDNA (∼14-fold) and nucDNA (∼10%; both p<0.01) that did not differ between 30-day survivors and non-survivors (**Figure 1A and 1B**). To corroborate these findings, we measured plasma levels of both mtDNA and nucDNA at multiple time points up to 4-hr post-ROSC in swine. Despite a brief ischemic period of insufficient duration to induce necrosis^20^, circulating mtDNA was markedly elevated (∼250-fold increase) at 1-hr post-ROSC (p<0.01) and remained ∼175-fold higher than baseline at 4-hr post-ROSC (p<0.01; **Figure 1C**). These increase in circulating mtDNA occurred despite a negligible rise in circulating nucDNA (**Figure 1D),** although total double-stranded DNA was also elevated at 1- and 4-hr post-ROSC (**Figure S3**).

**Figure 1.**
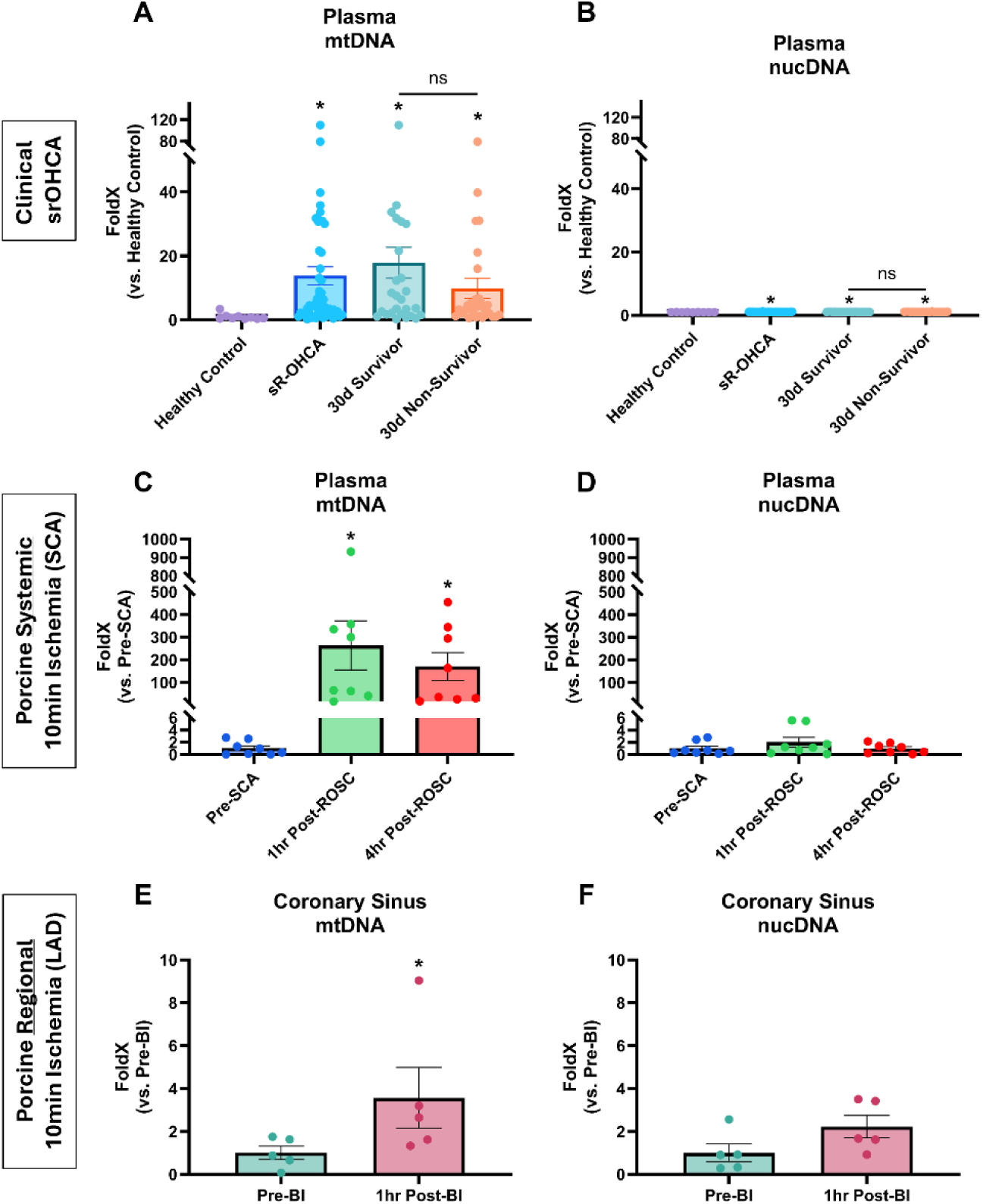
Selective Elevation of Circulating mtDNA Following SCA in Humans and Swine. qPCR analysis of 6hr post-ROSC whole plasma levels of A) mtDNA (Cyt-b) and B) nucDNA (β2M) within sR-OHCA patients stratified by 30-day survivorship and compared to health controls. n = 9, 57, 28, 29, respectively. qPCR analysis representing detected porcine circulating mtDNA and nucDNA before 10min untreated VF (Pre-SCA), 1-hr post-ROSC, and 4-hr post-ROSC, compared to a standard curve. C) mtDNA (Cyt-B) detection was normalized to isolated plasma volume, and fold-change was calculated compared to pre-SCA. D) nucDNA (β2M) detection was normalized to isolated plasma volume and fold-change was calculated compared to pre-SCA. Data represent biological n=8 and technical n=4. qPCR analysis representing detected porcine coronary sinus levels of mtDNA and nucDNA before (pre-BI) and 1-hr after reperfusion from regional brief ischemia (10-min) of the LAD perfusion territory, compared to a standard curve. E) mtDNA (Cyt-F) detection was normalized to isolated plasma volume, and fold-change was calculated compared to Pre-BI. D) nucDNA (β2M) detection was normalized to isolated plasma volume and fold-change was calculated compared to pre-BI. Data represent biological n=5 and technical n=4. Data represent mean ± SEM. *p < 0.01 vs. healthy controls/pre-SCA/pre-BI. NS = not significant. Intergroup relationships are highlighted by the presence of a bar.

Based on speculation that ischemic myocardium may be a primary source of mtDNA release, mtDNA and nucDNA levels were measured in the blood samples collected from the coronary sinus (CS) before and after brief myocardial ischemia induced by a 10-minute coronary occlusion. We have previously shown that this duration of regional myocardial ischemia causes a transient increase in myocyte TUNEL-positivity without pathologic evidence of myocardial necrosis^20^, while LAD occlusion of prolonged duration (≥15 minutes) is known to produce necrosis and infarction^29^. Brief ischemia (BI) led to a significant rise in CS levels of mtDNA (4.6 ± 1.4-fold vs. pre-BI; p<0.01) but not nucDNA (p=0.35) in the CS plasma 1-hr after reperfusion (**Figure 1E and 1F**). These data demonstrate that brief myocardial ischemia is associated with selective mtDNA release, even in the absence of necrosis. However, because the magnitude of mtDNA release after brief myocardial ischemia was dramatically lower than that observed after brief whole-body ischemia in swine with PCAS, extra-cardiac sources are likely a primary source of mtDNA after resuscitation from SCA.

Collectively, these clinical and pre-clinical data suggest that mtDNA is selectively released following brief ischemia, both systemically (SCA) and regionally (BI), which positions mtDNA as a potential pro-inflammatory stimulus to trigger leukocyte activation in PCAS.

### Intracellular mtDNA Increases Leukocyte Inflammatory Activation

We next explored whether elevated mtDNA (or nucDNA) drives innate immune cell responses *in vitro*. PBMCs were stimulated with either 2µg/mL of mtDNA or nucDNA, delivered in suspension or via transfection reagent (TR). Lipopolysaccharide (LPS) served as a positive control, and a TR alone condition served as a negative control. Only transfected mtDNA (mtDNA + TR) induced surface marker changes comparable to LPS stimulation, including increased expression of the inflammatory adhesion molecule (CD172+), and increased frequency of inflammatory dendritic cells (CD172+CD16+), granulocytes (CD172+CD163−), and macrophages (CD14+CD163+; **Figure 2A-2D**). In contrast, nucDNA, whether transfected or in suspension, did not provoke a similar inflammatory shift. Consistent with these phenotypic changes, mtDNA + TR stimulated significant secretion of TNFα (9.5±9.5 vs 402.0±38.9 pg/mL; p<0.05; **Figure 2E**), IL-1β (5.6±3.6 vs 46.8±6.9 pg/mL; p<0.05; **Figure 2F**), and IL-6 (0.0±0.0 vs 34.1±0.4 pg/mL; p<0.05; **Figure 2G**), as well as higher expression of inflammatory genes (NFκB, TNFα, IL-6, IFNα; **Figure S4**). Additionally, only mtDNA + TR stimulated a significant enrichment of intracellular levels of cGAMP, the endogenous second messenger of cGAS-STING (0.0±0.0 vs 2.7±0.4 pmol/mL lysate; p<0.05; **Figure 2H**). These findings indicate that leukocyte uptake of mtDNA is essential for triggering proinflammatory pathways, whereas nucDNA alone does not induce the same effect.

**Figure 2.**
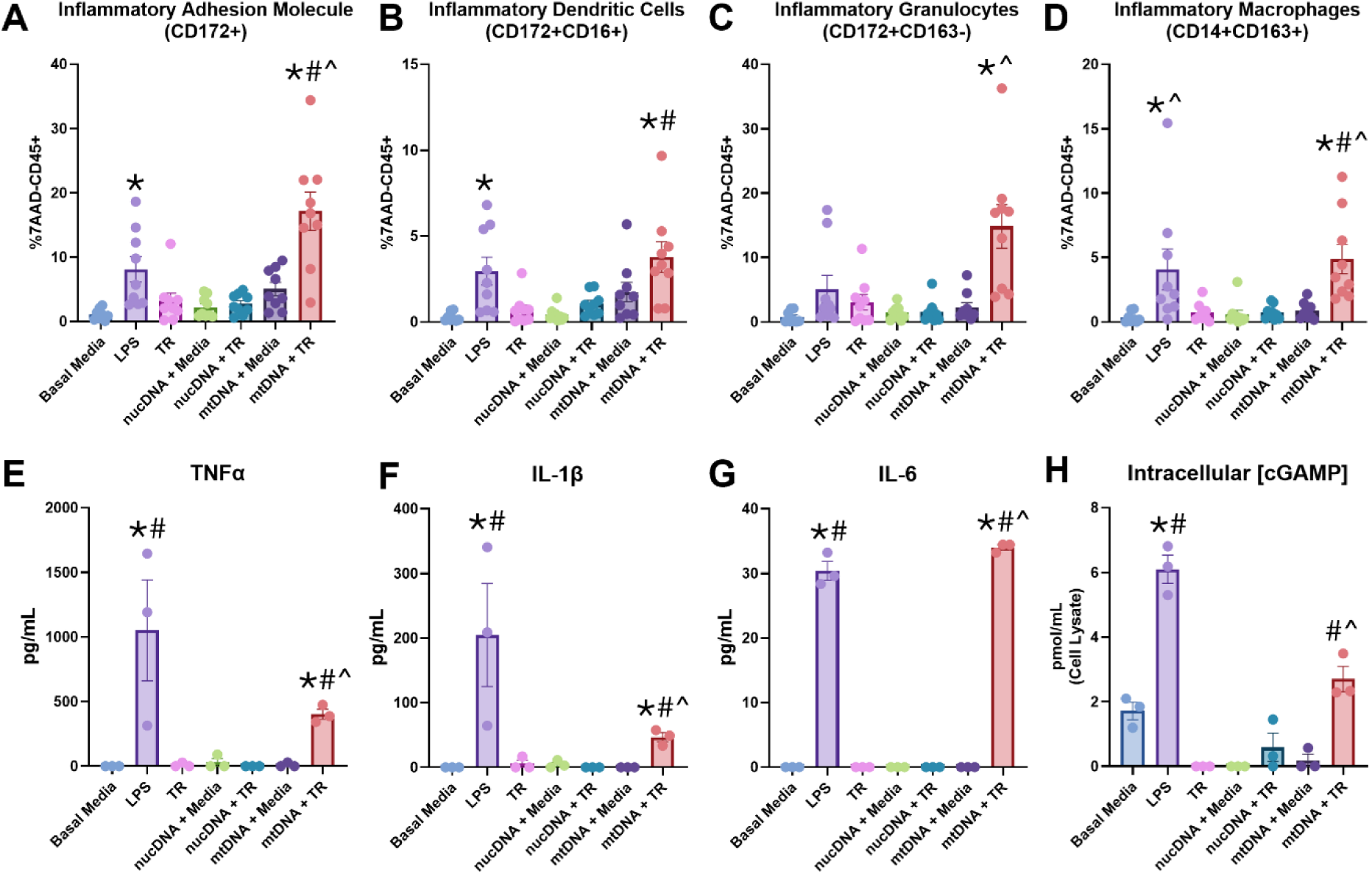
Intracellular mtDNA Increases Leukocyte Inflammatory Activation. Porcine PBMCs (CD45+7AAD-) were cultured for 24-hr in the presence of 2µg/mL nucDNA or 2µg/mL mtDNA in suspension (+media) or with lipofectamine 2000 (+TR). Flow cytometry was utilized to determine population proportion of A) cells expressing the inflammatory adhesion molecule CD172 (CD172+), B) inflammatory dendritic cells (CD172+CD16+), C) inflammatory granulocytes (CD172+163-), and D) inflammatory macrophages (CD14+CD163+). Following clarification, conditioned media levels of inflammatory cytokines E) TNFα, F) IL-1β, G) IL-6, and H) intracellular cGAMP levels were detected via ELISA. Data represent biological n=3 and technical n=3. Values are mean±SEM; *p< 0.05 vs. Basal Media. #p< 0.05 vs. TR-. ^p<0.05 vs. mtDNA+Media.

### Post-ROSC Plasma and mtDNA Stimulate Pro-Inflammatory Cell Expansion and Cytokine Release from Cultured PBMCs

To further assess whether mtDNA in post-ROSC plasma can shift leukocytes toward a pro-inflammatory state, PBMCs were cultured with plasma collected from swine 4-hr post-ROSC, mtDNA + TR, or LPS, compared with pre-SCA plasma, TR (–), or basal media, respectively. PBMCs exposed to mtDNA + TR displayed a surface marker profile that paralleled the effect of 4-hr post-ROSC plasma (**Figure 3A-3D**), including increased expression of CD172 and expansion of inflammatory cell subsets (dendritic cells, granulocytes, and macrophages). Like the effects seen following 4-hr post-ROSC plasma activation, mtDNA + TR stimulated a significant increase in secretion of TNFα (39.6±2.6 vs 310.1±9.9 pg/mL; p<0.05; **Figure 3E**) and IL-1β (65.3±23.2 vs 236.0±34.0 pg/mL; p<0.05; **Figure 3F**), compared to the TR (-) control. Together, these data suggest that mtDNA within post-ROSC plasma is capable of shifting leukocytes towards a pro-inflammatory phenotype and inducing inflammatory cytokine release.

**Figure 3.**
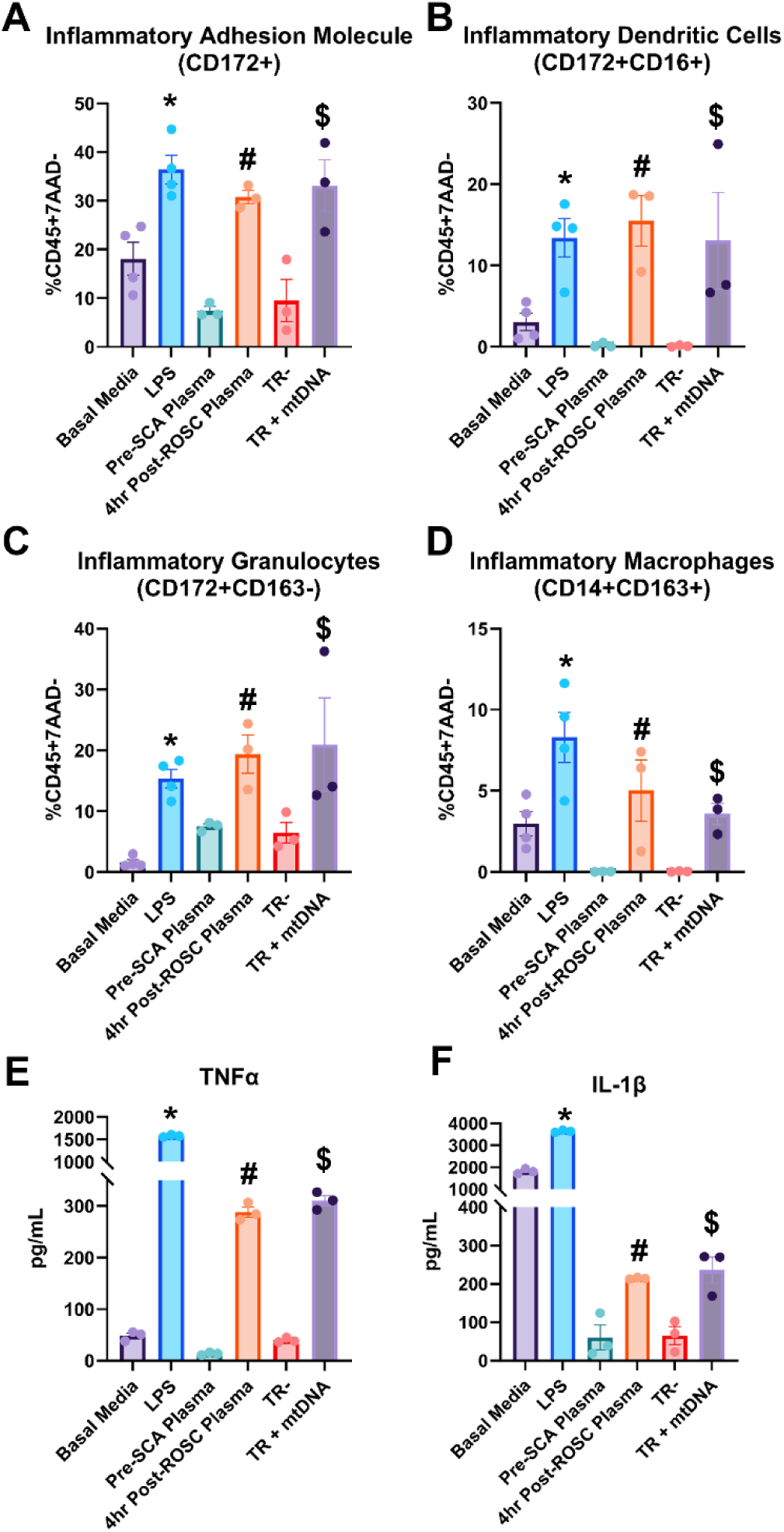
Post-ROSC Plasma and mtDNA Transfection Lead to Cultured PBMC Pro-Inflammatory Cell Differentiation and Cytokine Release. Porcine PBMCs (CD45+7AAD-) were cultured for 24-hr in the presence of LPS, 4-hr post-ROSC plasma, or 2µg/mL mtDNA with lipofectamine 2000 (TR+mtDNA), and compared to basal media, pre-SCA plasma, and TR – controls, respectively. Flow cytometry was utilized to determine population proportion of A) cells expressing the inflammatory adhesion molecule CD172 (CD172+), B) inflammatory dendritic cells (CD172+CD16+), C) inflammatory granulocytes (CD172+163-), and D) inflammatory macrophages (CD14+CD163+). Data represent biological n=4 and technical n=3. Following clarification, conditioned media levels of inflammatory cytokines E) TNFα and F) IL-1β were detected via ELISA. Data represent biological n=3 and technical n=3. Values are mean±SEM; *p<0.05 vs. Basal Media. #p<0.05 vs. pre-SCA Plasma. $p<0.05 vs. TR-.

### dsDNA is Required for Post-ROSC Plasma-Induced Immune Cell Activation

To determine whether mtDNA drives innate immune cell activation elicted by exposure to post-ROSC plasma, we selectively removed DNA and/or vesicles from 4-hr post-ROSC plasma. DNase I degraded free-floating DNA, whereas lysis buffer disrupted vesicles, and the combination of lysis and DNase I removed both vesicle integrity and dsDNA. As expected, 4-hr post-ROSC plasma induced expansion of inflammatory leukocyte sub-populations (CD172+ cells, macrophages, dendritic cells, granulocytes) that was significantly reduced following lysis or lysis + DNase I and rescued by reintroduction of mtDNA + TR (**Figure 4A–4D**). A similar pattern emerged for the secretion of TNFα and IL-1β (**Figure 4E-4F**) and for pro-inflammatory transcripts (NFκB, TNFα, IL-1α, IL-1β, IL-6, IL-8, IFNα, IFNβ; **Figure S5**). These results highlight the pro-inflammatory role of EV-encapsulated mtDNA and establish that mtDNA-containing EVs are required for post-ROSC plasma-induced inflammatory leukocyte activation.

**Figure 4.**
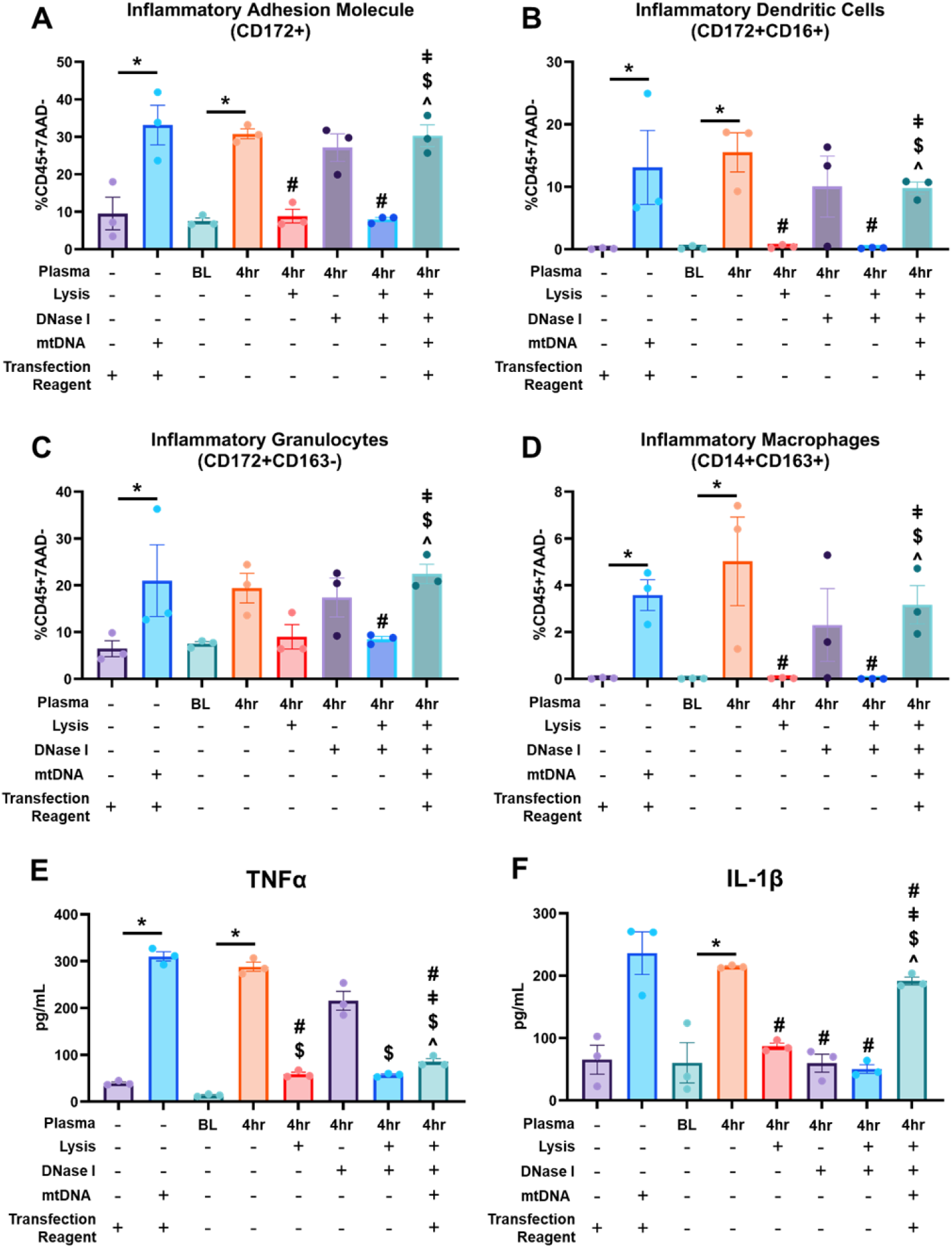
dsDNA is Required for Post-ROSC Plasma-Induced Immune Cell Activation. Porcine PBMCs (CD45+7AAD-) were cultured for 24-hr in the presence of 4-hr post-ROSC plasma, either in its native state or following treatment with lysis buffer to disrupt vesicles, DNase I to degrade dsDNA, or both. Rescue was achieved by the addition of 856ng mtDNA with lipofectamine 2000 (TR+mtDNA), as determined by 4-hr post-ROSC plasma mtDNA levels, in comparison to pre-SCA plasma (BL), and TR and mtDNA+TR controls. Flow cytometry was utilized to determine population proportion of A) cells expressing the inflammatory adhesion molecule CD172 (CD172+), B) inflammatory dendritic cells (CD172+CD16+), C) inflammatory granulocytes (CD172+163-), and D) inflammatory macrophages (CD14+CD163+). Following clarification, conditioned media levels of inflammatory cytokines E) TNFα and F) IL-1β were detected via ELISA. Data represent biological n=3 and technical n=3. Values are mean±SEM; *p<0.05 vs. BL plasma or TR (relationship indicated by bar). #p<0.05 vs. 4hr. $p<0.05 vs. 4hr + DNase I. ǂp<0.05 vs. 4hr + Lysis. ^p<0.05 vs. 4hr + Lysis + Dnase I.

### Post-ROSC Plasma-EVs Containing mtDNA Induce Inflammatory Leukocyte Activation

Given that EV disruption abrogated the immune response to post-ROSC plasma, we investigated whether plasma-EVs harboring mtDNA might be a primary driver of leukocyte activation. EVs were isolated from 4-hr post-ROSC plasma, with their identity confirmed by nanoparticle tracking (**Figure S6A-S6B**) and EV marker analysis (CD63, ANXA5, TSG101, Flot1, ICAM, ALIX, CD81; **Figure S6C-S6D**). Compared with pre-SCA EVs, 4-hr post-ROSC EVs contained significantly higher levels of D-Loop (∼300-fold; **Figure 5A**) and Cyt-B (∼300-fold; **Figure 5B**), with undetectable nucDNA levels (**Figure 5C**). In agreement with these preclinical data, EV-encapsulated mtDNA accounted for 79±3% of total circulating mtDNA in sR-OHCA patients compared with 55±7% in healthy controls (p<0.01; **Figure 6**). Interestingly, 30-day survivors (83±4%) exhibited significantly higher proportion of EV-mtDNA compared to 30-day non-survivors (74±5%, p=0.02; **Figure 6**). These results suggest that EV-mediated transfer of mtDNA to leukocytes may occur during the first several hours post-ROSC and implicate EV-encased mtDNA as a potential therapeutic target to attenuate post-ROSC leukocyte activation in PCAS.

**Figure 5.**
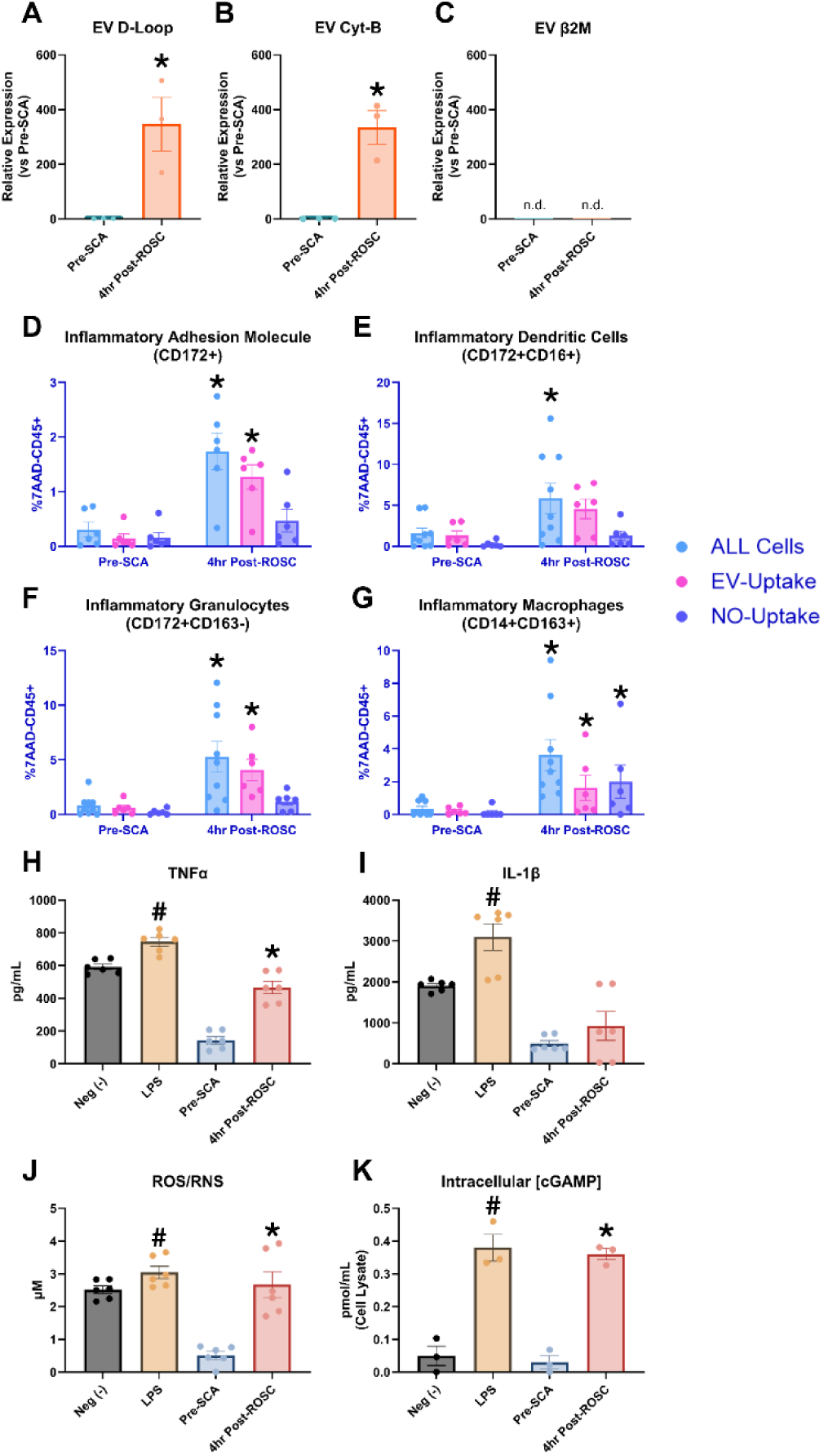
**Post-ROSC Plasma-EVs Containing mtDNA Induce Inflammatory Leukocyte Activation.** Porcine post-ROSC plasma-EVs were subjected to qPCR for detection of levels of mitochondrial genes A) D-Loop, B) Cyt-B, and C) nuclear β2M and expressed relative to their pre-SCA plasma-EV values. Data represent biologic n = 3. *p<0.05 vs. Pre-SCA. Porcine PBMCs (CD45+7AAD-) were cultured for 24-hr in the presence of 4-hr post-ROSC EVs (DiD stained) and were compared to pre-SCA plasma-EVs (DiD stained). Basal media (neg -) and LPS were used as controls. Flow cytometry was utilized to determine sub-population proportion of D) cells expressing the inflammatory adhesion molecule CD172 (CD172+), E) inflammatory dendritic cells (CD172+CD16+), F) inflammatory granulocytes (CD172+163-), and G) inflammatory macrophages (CD14+CD163+), while determining levels of all cells (blue), EV-uptake via DiD+ cells (pink), and No-EV-Uptake via DiD-cells (purple). Following clarification, conditioned media levels of inflammatory cytokines H) TNFα and I) IL-1β were detected via ELISA and J) levels of ROS/RNS were detected via DCF. K) Levels of intracellular cGAMP were detected via competitive ELISA. Data represent biological n=3 and technical n=2. Values are mean±SEM; #p<0.05 vs. neg (-). *p<0.05 vs. pre-SCA EVs. n.d. indicates non-detectible gene expression.

**Figure 6.**
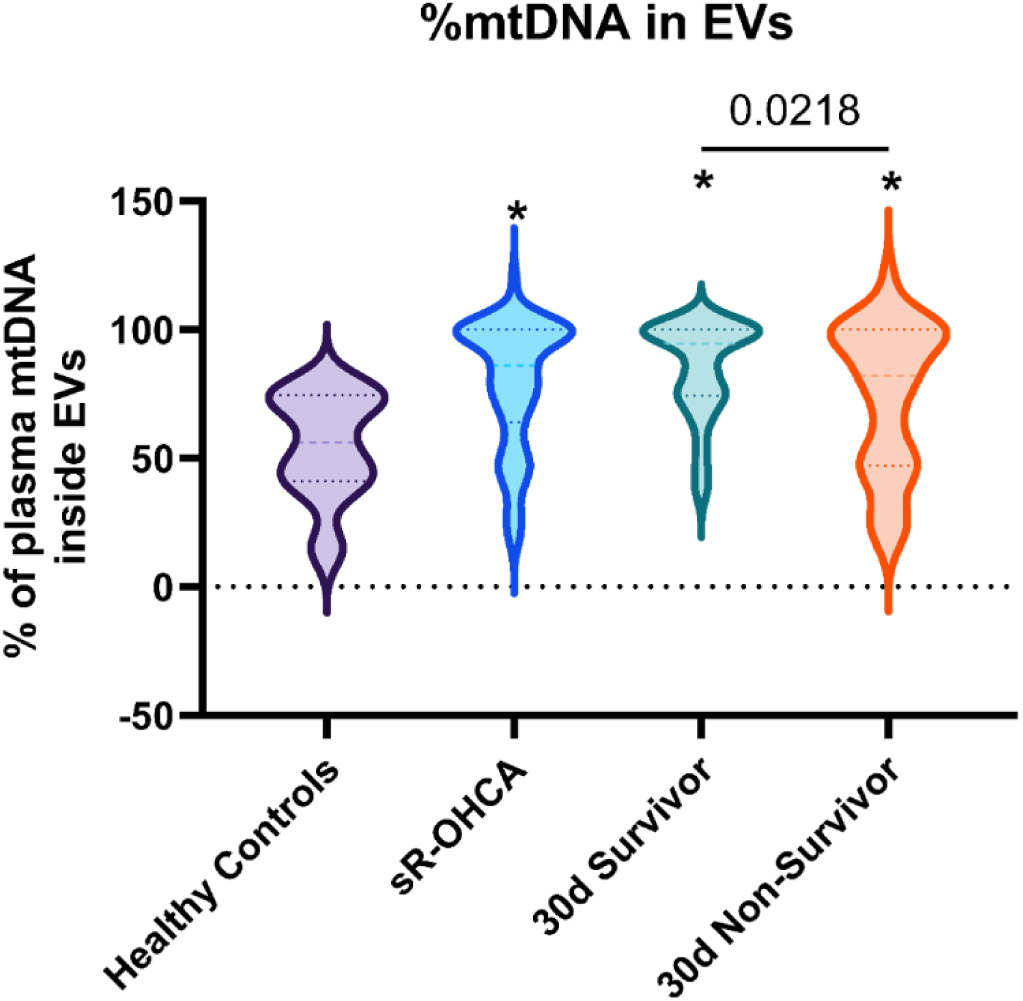
Elevated Proportion of Circulating EV-Encapsulated mtDNA in srOHCA Patients. qPCR analysis of plasma-EV levels of mtDNA (Cyt-b) were expressed as a fraction of DNA which was contained within plasma-EVs vs. whole plasma. n = 9, 57, 28, 29, respectively. *p < 0.01 vs. healthy controls. ns = not significant vs. healthy controls. Intergroup relationships are highlighted by the presence of a bar.

Since mtDNA is present within 4-hr post-ROSC plasma EVs and sr-OHCA plasma EVs, we postulated that EVs isolated from post-ROSC plasma could independently trigger immune cell activation. To test this hypothesis, we stimulated naïve PBMCs with DiD-labeled EVs isolated from 4-hr post-ROSC plasma and assessed changes in surface marker expression (**Fig S2, Bottom**) and inflammatory cytokine secretion. Compared to co-culture with EVs isolated from pre-SCA plasma, PBMCs which had taken up 4-hr post-ROSC plasma EVs (DiD+) exhibited a significant increase in inflammatory surface marker expression (**Figure 5D-5G).** Interestingly, post-ROSC plasma EVs elicited an increase in inflammatory macrophages that was not dependent on EV uptake (i.e., the expanded inflammatory macrophage population consisted of DiD+ and DiD-cells; **Fig 5G**). Compared to the effects of pre-SCA plasma EVs, post-ROSC plasma EVs induced a significant elevation of TNFα (144.7±22.6 vs 465.9±37.8 pg/mL; p<0.05; **Figure 5H**), IL-1β (495.5±75.7 vs 927.1±353.3 pg/mL; p<0.05; **Figure 5I**), and ROS/RNS (0.51±0.12 vs 2.67±0.39 µM; p<0.05; **Figure 5J**) within the conditioned media, along with significantly elevated intracellular cGAMP levels (0.03±0.02 vs. 0.36±0.02 pmol/mL lysate; p<0.05; **Figure 5K**). Together, these findings indicate that post-ROSC EVs containing mtDNA can independently trigger leukocyte activation when isolated from blood plasma.

### Post-ROSC Plasma EV–Encapsulated mtDNA Is Necessary for Inflammatory Leukocyte Activation

To verify that mtDNA within post-ROSC plasma EVs is essential for immune cell activation, we evaluated the effects of EV integrity disruption (lysis buffer) with and without simultaneous degradation of encapsulated DNA (lysis + DNase I) before PBMC stimulation. Following depletion and before PBMC stimulation, post-ROSC plasma EV integrity (**Figure S7A-S7B**) and dsDNA levels (**Figure S7C**) were confirmed to show expected treatment effects. The inflammatory phenotypes induced by 4-hr post-ROSC EVs were diminished by lysis buffer alone or lysis + DNase I, and were restored by reintroducing mtDNA + TR (**Figure 7A–7D**). Similarly, TNFα and IL-1β secretion (**Figure 7E-7F**), intracellular cGAMP levels (**Figure 7G**), and inflammatory gene expression (NFκB, TNFα, IL-1α, IL-1β, IL-6, IL-8, IFNα, and IFNβ; **Figure S8**) were blunted after EV disruption but partially recovered with mtDNA + TR. These results add further support to the notion that EV-encapsulated mtDNA within post-ROSC plasma is a key driver of leukocyte activation in PCAS.

**Figure 7.**
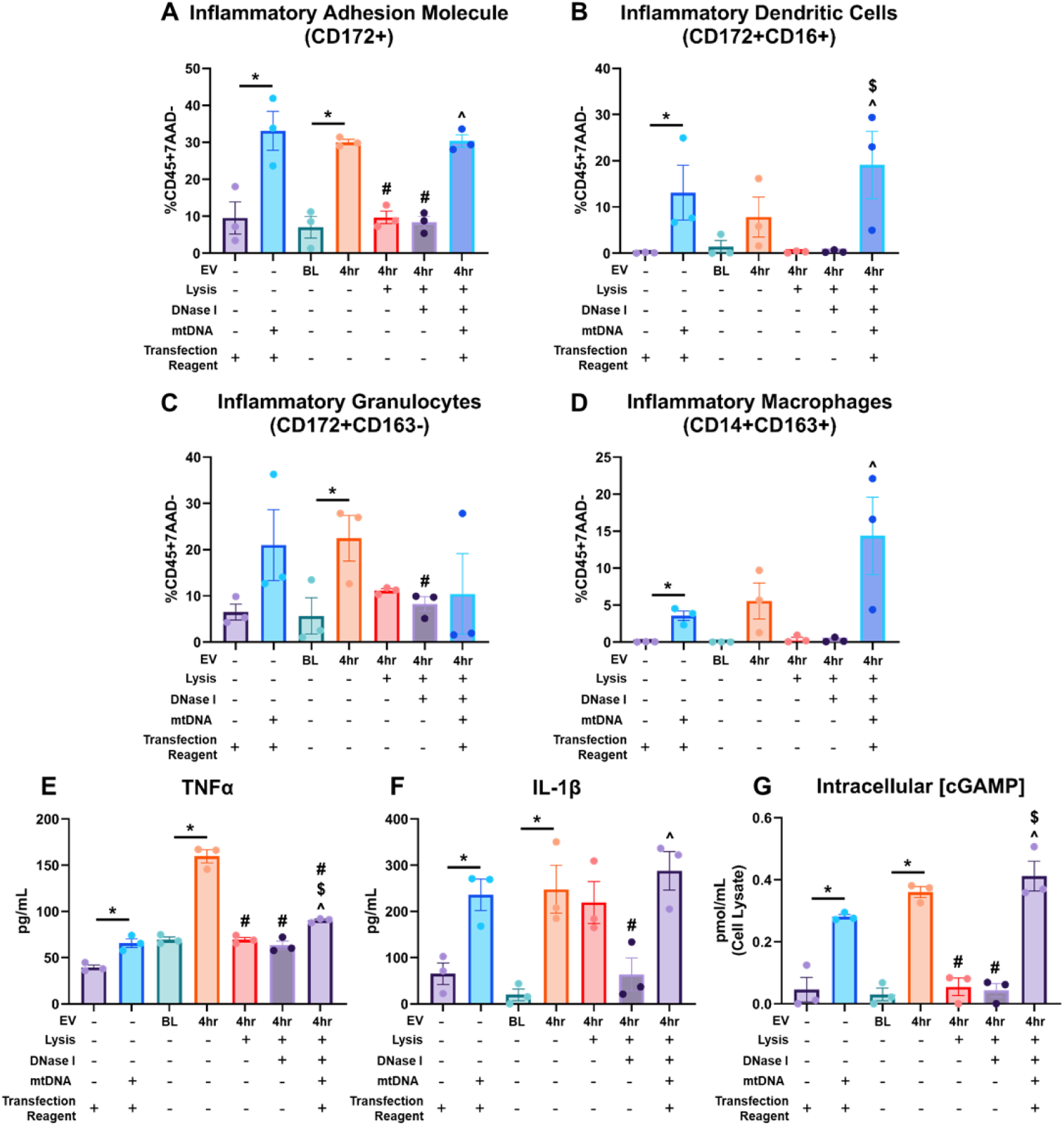
Post-ROSC Plasma-EVs Require mtDNA to Induce Inflammatory Leukocyte Activation. Porcine PBMCs (CD45+7AAD-) were cultured for 24-hr in the presence of 4-hr post-ROSC plasma EVs, either in their native state, treated with lysis buffer to disrupt vesicles, or treated with a combination of lysis buffer and DNase I to disrupt vesicles and degrade dsDNA. Rescue was achieved by the addition of 804ng mtDNA with lipofectamine 2000 (TR+mtDNA), as determined by 4-hr post-ROSC plasma EV mtDNA levels, in comparison to pre-SCA plasma EVs, and TR and mtDNA+TR controls. Flow cytometry was utilized to determine population proportion of A) cells expressing the inflammatory adhesion molecule CD172 (CD172+), B) inflammatory dendritic cells (CD172+CD16+), C) inflammatory granulocytes (CD172+163-), and D) inflammatory macrophages (CD14+CD163+). Following clarification, conditioned media levels of inflammatory cytokines E) TNFα, F) IL-1β, and intracellular cGAMP levels were detected via ELISA. Data represent biological n=3 and technical n=3. Values are mean±SEM; *p<0.05 vs. TR or BL plasma EVs (relationship indicated by bar). #p<0.05 vs. 4hr. $p<0.05 vs. 4hr + Lysis. ^p<0.05 vs. 4hr + Lysis + DNase I.

### Molecular Inhibition of TLR9 and cGAS Attenuates mtDNA-Induced Inflammation

Finally, to identify putative signaling pathways that could be targeted to attenuate mtDNA-mediated immune activation, we tested inhibitors targeting known dsDNA/mtDNA-sensing pathways: TLR9^17^ and cGAS^18^, as well as STING^30^ and PKG1^31^ downstream of cGAS. Transfection of mtDNA in the absence of inhibitors caused marked increases in inflammatory surface marker expression (**Figure 8A-8D and Figure S9A-S9D**), TNFα and IL-1β secretion (**Figure 8E-8F and Figure S9E-S9G**), and pro-inflammatory gene expression (**Figure S10 and Figure S9H-S9K**). In contrast, treatment with TLR9 antagonists (ODN 2088/inh-18), a cGAS antagonist (G140), a STING antagonist (H151), or a PKG1 antagonist (Rp-8-Br-PET-cGMP) significantly attenuated these responses in a concentration-dependent manner (**Tables S5-S7**). Likewise, intracellular levels of cGAMP were diminished following inhibition of TLR9 (ODN 2088), cGAS, STING, and PKG1 (**Figure 8G**), consistent with a mechanistic role of cGAMP within the signaling pathways driving mtDNA-induced immune activation. These results confirm that blocking mtDNA-sensing pathways effectively dampens the immune cell activation elicited by internalization of mtDNA.

**Figure 8.**
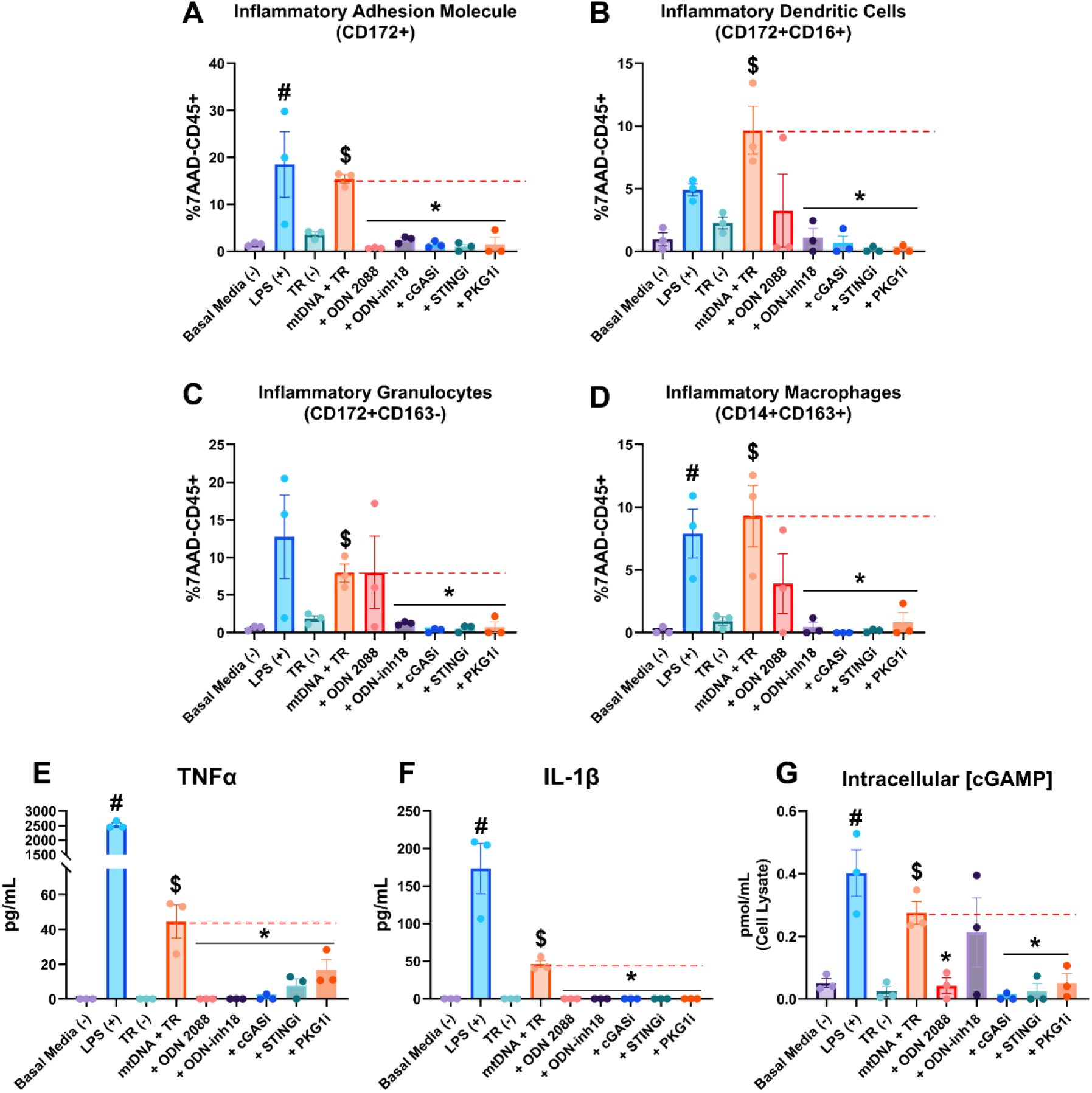
Molecular Inhibition of TLR9 or cGAS Signaling Pathways Attenuate mtDNA-Induced Leukocyte Inflammatory Activation. Porcine PBMCs (CD45+7AAD-) were cultured for 24-hr in the presence of LPS or 2µg/mL mtDNA with lipofectamine 2000 (TR+mtDNA) and compared to basal media (-) or TR – controls. At the time of stimulation, small molecular inhibitors were added against TLR9 (8µM ODN 2088 or 8µM ODN inh-18), cGAS (20µM G140), STING (4µM H151), and PKG1 (4µM Rp-8-Br-PET-cGMP). Flow cytometry was utilized to determine population proportion of A) cells expressing the inflammatory adhesion molecule CD172 (CD172+), B) inflammatory dendritic cells (CD172+CD16+), C) inflammatory granulocytes (CD172+163-), and D) inflammatory macrophages (CD14+CD163+). Data represent biological n=3 and technical n=3. Following clarification, conditioned media levels of inflammatory cytokines E) TNFα, F) IL-1β, and G) intracellular cGAMP levels were detected via ELISA. Data represent biological n=3 and technical n=3. Values are mean±SEM; #p<0.05 vs. Basal Media. $p<0.05 vs. TR-. *p<0.05 vs. mtDNA + TR (all conditions under bar are significant). Dashed red line indicates levels of mtDNA+TR in the absence of inhibitors.

## DISCUSSION

The present study provides several novel insights into how mtDNA drives post-resuscitation immune activation in PCAS. First, in sR-OHCA patients, circulating mtDNA levels were significantly elevated within the first several hours after ROSC in the absence of marked changes in circulating nucDNA levels, consistent with selective release of mtDNA in clinical PCAS. Second, in a porcine model of SCA, we detected a robust (∼250-fold) increase in mtDNA and negligible changes in nucDNA 1-hr post-ROSC, reaffirming the selective nature of mtDNA release even after a brief ischemic insult and in the absence of comorbid conditions. Third, *in vitro* experiments revealed that intracellular uptake of mtDNA, particularly when packaged within EVs, was required for inducing pro-inflammatory phenotypic shifts and cytokine secretion in cultured leukocytes, underscoring the functional significance of EV-encapsulated mtDNA. Fourth, pharmacological inhibition of TLR9 and cGAS-STING signaling notably dampened mtDNA-mediated leukocyte activation, suggesting that targeting these DNA-sensing pathways offers a viable strategy to curtail excessive inflammation. Collectively, these findings identify EV-encapsulated mtDNA as a novel mechanism by which immune cells become activated in PCAS and provide signaling pathways that warrant investigation as new therapeutic targets for interventions aiming to improve outcomes in patients resuscitated from SCA.

### Selective mtDNA Release and Immune System Activation

Previous studies have shown that cell-free dsDNA accumulates in the bloodstream after cardiac arrest, with some reports highlighting its prognostic value for neurological outcomes or survival^32–37^. However, these studies did not distinguish whether the source of dsDNA was mitochondrial or nuclear. Our results refine these observations by establishing that the substantial increase in dsDNA after SCA primarily reflects elevations in mtDNA. Furthermore, we demonstrated that EVs enriched with mtDNA make up a significant fraction of the dsDNA released following ROSC and is a key mediator of immune cell activation. This not only clarifies the underlying mechanism by which dsDNA might drive inflammation but also highlights the significance of mtDNA-loaded EVs in orchestrating the post-resuscitation inflammatory response.

The finding that mtDNA must be internalized by immune cells to incite a robust response aligns with prior work suggesting that mitochondrial components, particularly mtDNA, behave like bacterial DNA^38,39^. In our study, only transfected mtDNA (mtDNA + TR) elicited an inflammatory signature on par with LPS, whereas nucDNA did not. This underscores the unique ability of mtDNA to engage innate immune pathways, most notably TLR9 and cGAS-STING, which, once activated, prompt the production of TNFα, IL-1β, IL-6, and related pro-inflammatory mediators^39–44^.

### EV Encapsulation of mtDNA and Inflammatory Pathway Activation

By selectively removing DNA and/or vesicles from post-ROSC plasma, we found that EV integrity and encapsulated mtDNA are essential for its pro-inflammatory effects. This indicates that EVs are not merely passive carriers but actively determine the inflammatory potential of mtDNA by providing a mechanism for internalization by leukocytes. Similar conclusions emerge from prior studies linking mitochondria-derived vesicles (MDVs) and EVs to systemic inflammation, particularly in the context of trauma or infection^34–37,45,46^. Our data reinforce the concept that EV-encapsulated mitochondrial components, rather than free-floating mtDNA, drive leukocyte activation. Consistent with a “danger signal,” mtDNA in EVs can engage TLR9 and cGAS, eventually amplifying the innate immune cascade^39–42^.

The notion that post-ROSC EVs can stimulate macrophages, dendritic cells, and granulocytes also aligns with research showing how mtDNA-laden particles can propagate inflammatory signals beyond the cells initially stressed by ischemia/reperfusion injury^34,37^. Interestingly, inflammatory macrophage expansion involved both EV-positive (DiD+) and EV-negative (DiD–) cells, implying that once mtDNA-stimulated signaling is initiated, it may spread to bystander immune cells, further enhancing systemic inflammation in PCAS.

### Implications for Clinical Outcomes in PCAS

The “sterile sepsis”–like state that often arises after OHCA has been attributed to overactive immune responses^38,47^. Our findings suggest that mtDNA release, specifically mtDNA encapsulated in EVs and released into plasma, is a key contributor to this phenomenon. This perspective resonates with observational data linking high inflammatory biomarker levels to circulatory failure, organ dysfunction, and increased mortality after cardiac arrest^48^. Moreover, although some studies have proposed that dsDNA or dsDNA/DNase ratios correlate with patient prognosis^32,33^, the presence of EV-encapsulated mtDNA may complicate the predictive power of these measurements, since DNase has limited capacity to degrade mtDNA that is encased within vesicles.

In our cohort of sR-OHCA patients, we found that the portion of circulating mtDNA within EVs was significantly higher than in healthy controls. Notably, we also observed that 30-day non-survivors exhibited a lower proportion of EV-encapsulated mtDNA compared with 30-day survivors at the early post-ROSC timepoint. EV uptake can occur rapidly, with EVs being identified inside cells from as early as 15 minutes following exposure to the target cell^49^. Therefore, it could be hypothesized that rapid cellular uptake of EV-mtDNA in the non-survivor group may have led to an accelerated and/or more severe immune activation, whereas higher residual levels of EV-encapsulated mtDNA in 30-day survivors suggest a different kinetic pattern of release and uptake. Further investigation of post-ROSC EV kinetics, including timing, biodistribution, and cellular internalization, will be essential for deciphering how these differences may be related to patient outcomes and exploring whether combining measures of total dsDNA, nucDNA, and EV-encapsulated mtDNA could refine risk stratification or treatment in PCAS.

### Targeting mtDNA-Sensing Pathways

We observed that antagonizing TLR9 and cGAS-STING significantly diminished intracellular levels of cGAMP and inflammatory responses induced by mtDNA. This bolsters evidence that these DNA-sensing pathways are pivotal in orchestrating the post-ROSC inflammatory milieu and suggests new therapeutic angles for modulating immune activation^39–44^. Although our study investigated TLR9 and cGAS-STING, other cytoplasmic DNA sensors like Rad50 and Ku70 may also respond to mtDNA release, potentially contributing to the innate immune activation in PCAS^44,50^. Broadening future investigation to include these sensors could uncover additional molecular targets to mitigate PCAS-associated inflammation.

### Future Directions

The precise mechanisms by which mtDNA is packaged into EVs or shuttled via MDVs following ROSC remains incompletely understood, but it is plausible that oxidative stress, mitochondrial fission, and/or disrupted autophagic flux underlie this phenomenon^45,46^. In parallel, the contributions of distinct immune cell subsets, diverse tissue sources, and the kinetic profile of EV-mediated mtDNA release to systemic inflammation have yet to be systematically characterized. Pharmacologic approaches aimed at blocking mtDNA liberation, inactivating vesicle-associated mtDNA, and/or inhibiting critical DNA-sensing pathways hold promise for alleviating the inflammatory burden of PCAS. Validating these strategies in large animal models and subsequent clinical trials will be critical to determining their efficacy in mitigating mtDNA-driven injury.

Additionally, it is still unclear why some individuals exhibit more pronounced mtDNA release or a more severe inflammatory response post-ROSC. Heterogeneity in mitochondrial function, redox balance, or DNase activity may contribute to varying extents of mtDNA release. Studies also suggest that partial EV loading of mtDNA might occur under more subtle damage conditions, while severe cell death could release substantial amounts of free mtDNA^37,45^. Clarifying these distinct mechanisms could enable refined therapeutic interventions that target harmful mtDNA release without compromising vital metabolic and/or immunological functions.

## Limitations

Although the porcine model of PCAS exhibits key features of the clinical syndrome, inter-species differences in physiology and disease course compared with human subjects should be considered when interpreting preclinical data. The partial translation of our findings in swine to human sR-OHCA patients provides initial clinical support; however, larger cohorts with extended follow-up are needed to study a potential link between mtDNA-EVs and clinical outcomes. We also did not explore other DNA-sensing pathways beyond TLR9 and cGAS-STING, leaving room to investigate additional innate immune DAMP receptors which might significantly influence the post-ROSC inflammatory response.

## Conclusion

In summary, our findings identify a novel mechanism of selective mtDNA release following brief whole-body ischemia and implicate EV-encapsulated mtDNA as a central mediator of sterile inflammation in PCAS. By demonstrating that mtDNA release and vesicular packaging are indispensable for TLR9 and cGAS-STING driven immune activation, we unveil a novel mechanism by which an excessive inflammatory response may exacerbate organ damage following SCA. These new insights fuel interest in evaluating the therapeutic potential of strategies targeting DNA-sensing pathways to lessen the excessive inflammatory burden that accompanies resuscitation. Pharmacological disruption of mtDNA-mediated immune activation, whether by inhibition of its release, vesicular encapsulation, and/or downstream receptor signaling, holds promise for improving outcomes in the large group of patients who develop PCAS following resuscitation from SCA.

## Data Availability

The data supporting the study's findings are available from the corresponding author upon reasonable request. Please see Supplemental Materials for complete methodological details.

## Acknowledgments

This project was made possible with support from the Laboratory Animal Facilities at the State University of New York at Buffalo, the Confocal Microscope and Flow Cytometry Facility at the State University of New York at Buffalo, and the Flow and Image Cytometry Shared Resource at Roswell Park Comprehensive Cancer Center. We are grateful for the excellent technical support provided by Elaine Granica, Maggie Vogel-Cryan, LVT, Rebeccah Young, MA, and Beth Palka, without whom these studies could not have been completed.

## Author Contributions

Acquisition of funding: TJR, BRW; conceptualization: TJR, BRW; methodology: TJR, BRW; data analysis, TJR, BRW; investigation, TJR, ERH, SZ, DC, LG, BRW; preparation of initial manuscript, TJR; manuscript review and editing, TJR, ERH, SZ, DC, LG, BRW. All authors have read and agreed to the final version of the manuscript.

## Supplemental Materials

Supplemental Materials Extended Methods Supplemental Figure S1-S10 Supplemental Tables S1-S7

## Non-Standard Abbreviations and Acronyms

AMI: Acute Myocardial Infarction
ANOVA: analysis of variance
BI: brief ischemia
CM: conditioned media
CPR: cardiopulmonary resuscitation
dsDNA: double stranded DNA
DAMP: danger associated molecular pattern
EPI: epinephrine
EV: extracellular vesicle
FBS: fetal bovine serum
IQR: interquartile range
LAD: left anterior descending
LPS: lipopolysaccharide
MDV: mitochondria-derived vesicle
tDNA: mitochondrial
DNA nucDNA: nuclear DNA
OHCA: out-of-hospital cardiac arrest
PBMCs: peripheral blood mononuclear cells
PBS: phosphate buffered saline
PCAS: post cardiac arrest syndrome
ROS/RNS: reactive oxygen species/reactive nitrogen species
ROSC: return of spontaneous circulation
SBP: systolic blood pressur
SCA: sudden cardiac arrest
SEM: standard error of the mean
sR-OHCA: successfully resuscitated out-of-hospital cardiac arrest
TR: Transfection Reagent
TTM: targeted temperature management
VF: ventricular fibrillation
WBC: white blood cell

